# Gut Health, Nutrient intake and Well-being in Community-Dwelling Older Adults

**DOI:** 10.1101/2019.12.16.19015008

**Authors:** F. Fart, L. Tingö, S. Engelheart, C. M. Lindqvist, R. J. Brummer, A. Kihlgren, I. Schoultz

## Abstract

**Background:** A majority of community-dwelling older people will in the near future be in need of increased health care. By investigating the relationship between gut health, well-being and nutrient intake we aim to recognize areas through which health might be promoted.

**Methods:** A cross-sectional observational study was performed enrolling 229 older adults (≥65 years). Validated questionnaires were used to assess gut health, nutrient intake and well-being.

**Results:** 65% of the participant experienced gastrointestinal symptoms. Gastrointestinal symptoms significantly correlated to anxiety, stress and decreased quality of life. Dyspepsia correlated to a lower energy percentage of protein. An intake below the nutritional recommendations was found for protein, fibre, fat (monounsaturated/polyunsaturated), while an intake above the recommendations was found for saturated fats and alcohol

**Discussion:** A majority of the community-dwelling older adults experienced gastrointestinal symptoms and had an imbalanced macronutrient intake. Gut health, diet and well-being all represents important areas for future intervention studies.

## Introduction

In the last decades, lifespan has increased dramatically due to improved health and longevity, leading to a global ageing phenomenon (Cohen, 2003; World Health Organisation, 2002). The increase of the aged population will have an impact on health-care systems and socio-economics worldwide due to increased prevalence of age-related diseases and hospitalisations. As the life expectancy of the population increases, there is a growing awareness of the importance of promoting optimal functionality and health throughout life.

A well-functioning gastrointestinal (GI) tract has been identified as essential for health and well-being among older adults (Algilani et al., 2014). Diseases of the GI tract, including inflammatory bowel disease and irritable bowel syndrome, are known to lead to a negative health status in older adults (El-Serag, Olden, & Bjorkman, 2002; J.-P. Ganda Mall et al., 2018; Knowles et al., 2018). These data emphasise gut health as a potential area through which health and well-being might be promoted.

Dietary intake and dietary patterns are known to affect health and well-being (Jacka et al., 2010) and also known to influence gut health either through direct effects (J. P. Ganda Mall et al., 2018) or indirectly by influencing the gut microbiota composition (Claesson et al., 2012; Mitsou et al., 2017). Age-related physiological changes are further known to influence older adults’ nutrient intake, such as decreased sense of smell, taste, satiety and hunger (S. B. Roberts & Rosenberg, 2006). Thus, it is essential to analyse nutrient intake in relation to GI health.

A relationship between nutrient intake, intestinal microbiota and health status has previously been reported among older adults (Claesson et al., 2012). However, the prevalence of GI symptoms among older adults living at home has not been thoroughly elucidated. This is an essential group to include when investigating healthy ageing, as the majority of these individuals will need elevated health care resources in the near future. In the present study, we aim to investigate the connection between gut health, well-being and nutrient intake in order to open up for further approaches targeting gut health to increase well-being and independence among older adults.

## Methods

### Study participants

Participants were recruited by advertisements in local newspapers and in local senior residences in Örebro Sweden. In total, 302 participants were assessed for eligibility, of these 229 were included in the final analyses, figure 1. Inclusion criterion was: age ≥ 65 years.

**Figure 1.**
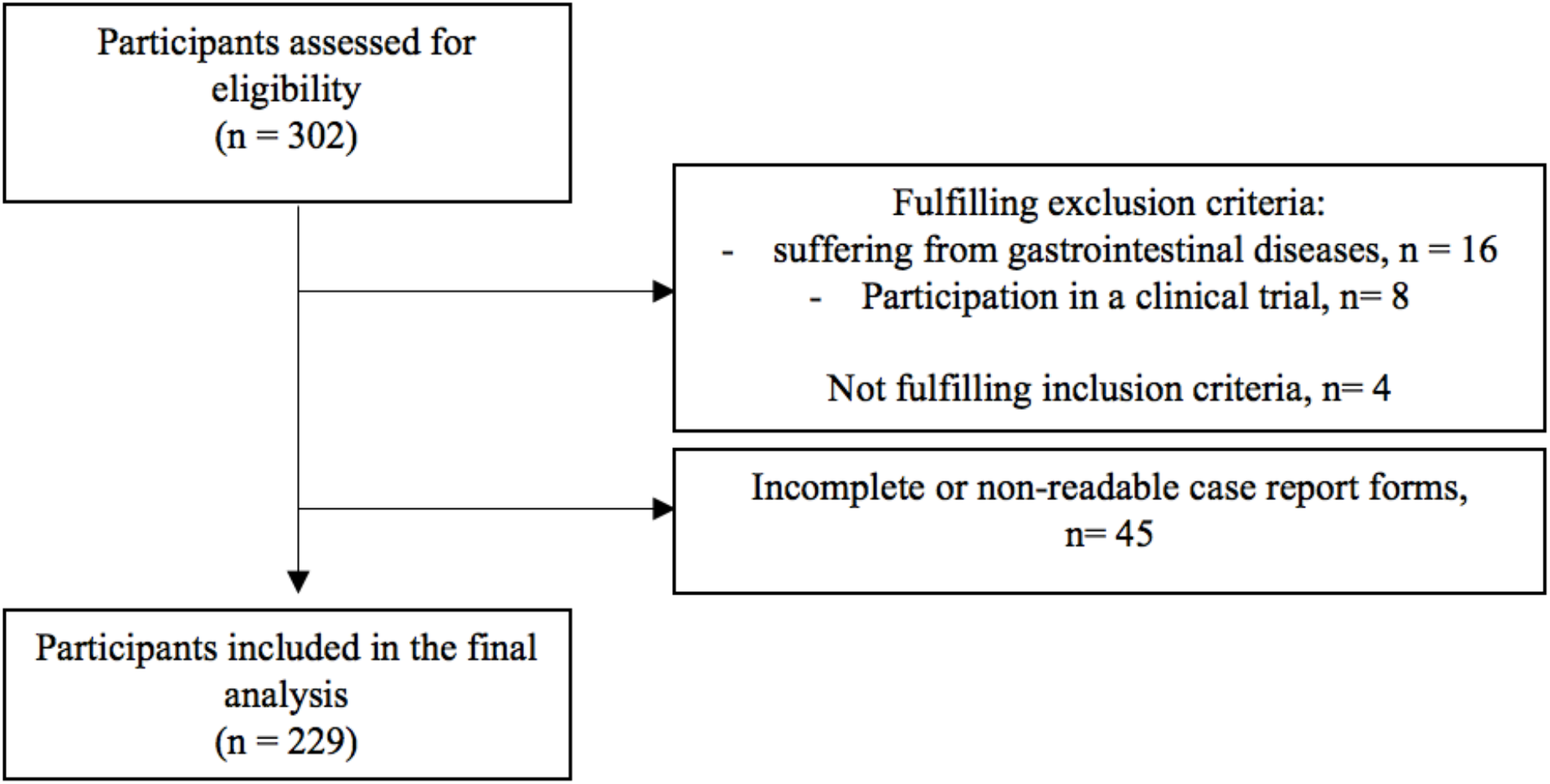
Participant flow illustrating the inclusion process of study participants

Exclusion criteria included: any known gastrointestinal disease with strictures, malignancies or ischemia, including **i**nflammatory bowel disease; participation in other clinical trials in the past three months.

### Data collection

All data sampling was conducted in the participants’ own home environment with support of a contact person, i.e. a medical student informed in detail about the study procedures. All demographic data were recorded in the case report form (CRF), except physical activity, which was measured by the validated questionnaire Frändin-Grimby Activity Scale (FGAS) (Frändin & Grimby, 1994). The instrument evaluates the respondents’ activity level during summer and winter using fixed response alternatives.

The experience of gastrointestinal symptoms was assessed using the Gastrointestinal Symptoms Rating Scale (GSRS). It is a validated instrument and has previously been used in an older population (Svedlund, Sjodin, & Dotevall, 1988). The scale measures 15 symptoms, which are divided into five symptom domains: reflux, abdominal pain, dyspepsia, diarrhoea and constipation. Each domain was scored individually to evaluate the prevalence of each symptom, while a total GSRS score (the mean score of all five symptom domains) was used to estimate the overall gastrointestinal discomfort. A score >2 was considered as having a GI symptom.

The nutrient intake was estimated by a semi-quantitative Food Frequency Questionnaire (FFQ) asking for dietary intake the past year. The method has been previously validated and described by Johansson I et al (Johansson et al., 2002). Participants estimated their intake of 66 food items from 0-8 (0=never, 8=4 or more times a day). To facilitate inter-individual comparisons, the intake per day was expressed as energy percentage (E%) and intake of fibre was expressed as gram per MJ energy intake. Dietary intakes were compared to the Nordic Nutritional Recommendation (NNR)(*Nordic Nutrition Recommendations 2012* : *integrating nutrition and physical activity*, 2014). In addition data regarding health-related quality of life (HRQOL) was assessed through the EuroQol (EQ) (Rabin & de Charro, 2001). Psychological distress and the experience of stress was estimated through the validated Hospital Anxiety and Depression Scale (HADS) (M. H. Roberts, Fletcher, & Merrick, 2014) and the Perceived Stress Scale (PSS) (Ezzati et al., 2014), respectively. Outlined in more detail in table S1 in appendix.

### Data analysis and statistics

Median values, with the interquartile range (IQR), were generated for all demographic and questionnaire data. Missing data of less than two items per questionnaire (except FFQ) was imputed by the arithmetic mean or according to the instruction for the specific questionnaire. In total 29 individual items were imputed (FGAS: 8, GSRS: 7, HADS: 4, PSS: 10). Forty individual questionnaire sheets had to be excluded from the analysis due to missing values in proportions that did not allow for imputation. An additional 7 sub-variables on EQ-index and GSRS could not be imputed. Stratifications were performed for: sex (male/female) and GI symptom as judged by a score >2 on the total score of GSRS (yes/no) and analysed with Mann-Whitney U test and Chi-square test. For comparison between the parameters, a Spearman correlation using Bonferroni correction for multiple comparisons was performed. All statistical analysis was performed using SPSS version 24 for Mac (SPSS software, IBM corporation, USA), p<0.05 or q<0.05 were considered significant.

### Ethical considerations

This study was conducted according to the guidelines of the Declaration of Helsinki and was approved by the Regional Research Ethics Committee in Uppsala, Sweden (Dnr: 2012/309).

## Results

### Demographic data

All demographic data are presented in table 1. The median age of the study population was 71 years whereof 67% were females. Being a current smoker was reported by 5% of the study population. Polypharmacy, taking five or more medications, were reported by 18%.

**Table 1.** Participants characteristics

| **Parameter** | **All participants,**<br>**n=229** | **Males**<br>**n= 72** | **Females**<br>**n= 157** | **p-value** |
| --- | --- | --- | --- | --- |
| **Age** |  |  |  |  |
| Median (IQR) | 71 (68 - 75.5) | 72 (69 - 76) | 71 (68 - 75) | 0.311 |
| **Higher education** |  |  |  |  |
| N (%) | 96 (43%) | 36 (49%) | 60 (40%) | 0.204 |
| **Smoking** |  |  |  |  |
| N (%) | 11 (5%) | 3 (4%) | 8 (5%) | 1.0 |
| **Physical activity** |  |  |  |  |
| Median (IQR) | 3.5 (3 - 4) | 3.5 (3 - 4) | 3.5 (3 - 4) | 0.613 |
| **Polypharmacy** |  |  |  |  |
| N (%) | 39 (18%) | 11 (16%) | 28 (19%) | 0.595 |
| **Number of medicines** |  |  |  |  |
| Median (IQR) | 2 (1 - 4) | 2 (1 - 3) | 2 (1 - 4) | 0.457 |
| **Retirement age** |  |  |  |  |
| Median (IQR) | 65 (63 – 65) | 65 (63 - 67) | 65 (62 - 65)* | **0.017** |
| **Have a partner** |  |  |  |  |
| N (%) | 127 (55%) | 54 (72%) | 73 (47%)* | **<0.001** |
| **GI symptoms** |  |  |  |  |
| Median (IQR) |  |  |  |  |
| Dyspepsia | 2.0 (1.5 - 3.0) | 2.0 (1.3 - 2.9) | 2.1 (1.5 - 3.3) | 0.258 |
| Constipation | 1.7 (1.0 - 2.7) | 1.3 (1.0 - 2.3) | 1.7 (1.3 - 3.0)* | **0.021** |
| Abdominal pain | 1.3 (1.0 - 2.0) | 1.3 (1.0 - 1.8) | 1.7 (1.0 - 2.3)* | **0.023** |
| Diarrhoea | 1.3 (1.0 - 2.3) | 1.3 (1.0 - 2.3) | 1.3 (1.0 - 2.3) | 0.328 |
| Reflux | 1.0 (1.0 - 1.5) | 1.0 (1.0 - 1.5) | 1.0 (1.0 - 2.0) | 0.952 |
| **HADS-depression** |  |  |  |  |
| Median (IQR) | 2 (1 - 3) | 3 (1 - 5) | 1 (1 - 3)* | **0.041** |
| **HADS-anxiety** |  |  |  |  |
| Median (IQR) | 3 (1 - 5) | 3 (1 - 5) | 3 (1 - 5) | 0.395 |
| **PSS-stress** |  |  |  |  |
| Median (IQR) | 10 (6 - 14) | 10 (6 - 14) | 10 (6 - 15) | 0.632 |
| **EuroQol** |  |  |  |  |
| Median (IQR) |  |  |  |  |
| Index | 0.8 (0.8 - 1.0) | 0.9 (0.8 - 1.0) | 0.8 (0.8 - 0.9) | 0.059 |
| VAS | 80 (75 - 90) | 80 (75 - 90) | 80 (70 - 90) | 0.732 |

\* $p < 0.05$ were considered significant.

### Gastrointestinal symptoms

In total 65% of the study population experienced one or several GI symptoms, whereof 5% had severe symptoms. For each symptom category, the following prevalence was reported: 48% for dyspepsia, 33% constipation, 26% abdominal pain, 23% diarrhoea and 15% reflux. The majority of the study population experienced more than one symptom, median 2 (IQR: 1-3), as illustrated in figure S1 in appendix. Females scored significantly higher (i.e. experienced more severe symptoms) than males on the subdomains for constipation and abdominal pain (table 1).

### Nutrient intake

The nutrient intakes for the study participants are shown in table 2. The median total energy intake was found to be 5.5 MJ/day (i.e. 1314 kcal/day), where males had a significantly higher intake compared to females. The intake of several macronutrients did not meet the NNR as illustrated in figure 2. Males had higher E% of fat and saturated fat intake, but a lower E% of carbohydrate, protein and intake of fibre (g/MJ) than females. A higher proportion of males than females ate less than the NNR of E% of protein and fibre g/MJ. Having GI symptoms were associated with a low E% protein intake (p=0.003), a higher E% from carbohydrates (p=0.019) and higher estimated E% of added sugars (p=0.003) (see table s2 in appendix).

**Table 2.** Stratifications for macronutrient

| **Macronutrient intake** | **Nordic Nutrition Recommendation** | **All** | **Males** | **Females** | **p-value** |
| --- | --- | --- | --- | --- | --- |
| **Total energy intake, MJ/day<sup>1</sup>** | Male: 6.1 | 5.5 | 7.9 | 4.9* | **<0.001** |
| Median (IQR) | Female: 5.0 | (4.5 - 7.3) | (6.2 - 10.0) | (4.2 - 5.8) |  |
| **Protein, E%** | 15-20 E% | 14.6 | 13.9 | 15.2* | **0.001** |
| Median (IQR) |  | (12.8 - 16.6) | (12.3 - 15.6) | (13.2 - 16.8) |  |
| **Fibre, g/MJ** | >3g/MJ | 2.3 | 2.7 | 3.0* | **0.002** |
| Median (IQR) |  | (1.9 - 2.7) | (2.2 - 3.2) | (2.5 - 3.5) |  |
| **Fat, E%** | 25-40 E% | 34.0 | 36.1 | 33.3* | **0.004** |
| Median (IQR) |  | (30.5 - 37.8) | (33.4 - 40.2) | (29.5 - 36.9) |  |
| **Saturated fat, E%** | <10 E% | 14.5 | 15.8 | 14.1* | **0.022** |
| Median (IQR) |  | (12.8 - 17.1) | (13.6 - 18.2) | (12.2 - 16.8) |  |
| **Monounsaturated fat,** | 10-20 E% | 11.0 | 10.8 | 11.1 | **0.158** |
| **E%** |  | (9.6 - 12.3) | (9.5 - 12) | (9.6 - 12.3) |  |
| Median (IQR) |  |  |  |  |  |
| **Polyunsaturated fat,** |  |  |  |  |  |
| **E%** | 5-10 E% | 5.0 | 4.9 | 4.9 | 0.684 |
| Median (IQR) |  | (4.0 - 6.2) | (3.7 - 6.1) | (4.0 - 6.1) |  |
| **Carbohydrates, E%** | 45-60 E% | 48.0 | 45.2 | 48.8* | **0.029** |
| Median (IQR) |  | (42.7 - 51.7) | (40.2 - 49.2) | (44.0 - 52.6) |  |
| **Estimated added** |  |  |  |  |  |
| **sugars, E%** | <10 E% | 4.7 | 4.5 | 4.7 | 0.724 |
| Median (IQR) |  | (3.4 - 6.2) | (3.2 - 6.6) | (3.6 - 6.2) |  |
| **Alcohol, E%** | <5 E% | 2.5 | 3.0 | 2.3 | 0.121 |
| Median (IQR) |  | (1.2 - 5.2) | (1.3 - 5.6) | (1.1 - 4.8) |  |

MJ – megajoule, IQR – interquartile range, E% - energy percentage. \*p<0.05 were considered significant.
<sup>1</sup>Estimated resting metabolic rate of older adults.

**Figure 2:**
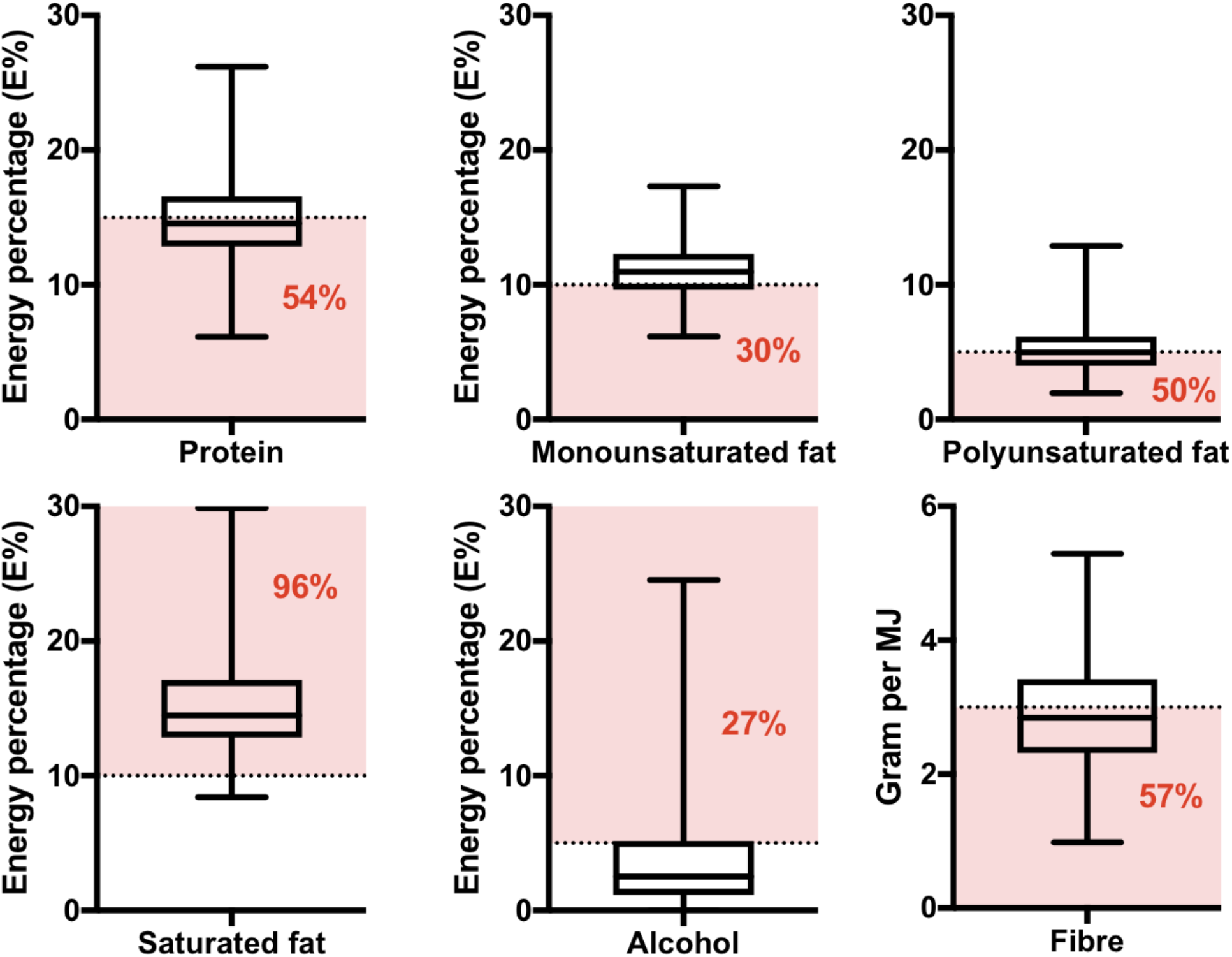
Schematic overview of the intake of macronutrients (protein, mono-/polyunsaturated and saturated fats, alcohol and fibre) in the study population in correspondence to the Nordic Nutritional Recommendation (NNR). The macronutrients are presented as energy percentage (E%), where the dotted line represents the minimal recommended intake level. The percentage in red and the coloured box (pink) indicates the proportion of the study participants having an intake below or above the recommended level. All data are presented as E% in box plots showing median, interquartile range (IQR) for each macronutrient.

### Well-being

The majority of the study population scored above 75% of the optimal score on questionnaires depicting subjective well-being (quality of life, anxiety, depression and stress). According to HADS cut-off (>7), 4% and 9% of the participants reported a score categorised as borderline for depression and anxiety respectively. Males reported higher scores for depression (p=0.041) and 11% of the male participants were categorised as being on the borderline for depression compared to 1% of the female participants (p=0.002). Those with a GI symptom had lower HRQOL as indicated by EuroQol (index, p=0.001, and VAS p=0.002). GI symptoms were also associated with higher scores for anxiety (p=0.003) and stress (p=0.006).

### Correlations

Significant correlations were found between mean GSRS score and anxiety (r=0.303, q<0.001), stress (r=0.233, q=0.009) and decreased HRQOL (index and VAS-scale, r=-0.394 and r=-0.345 respectively, q<0.001). Similar patterns could be seen for the individual subdomains, except for reflux, which did not affect well-being. Dyspepsia was found to correlate to a low E% of protein intake (r= -0.199, q=0.031). A trend was found between dyspepsia and a high intake of estimated added sugars (r=0.181, q=0.055). Moreover, physical activity correlated significantly to stress (r=-0.195, q=0.031), HRQOL (VAS, r=0.297, q<0.001; index r=0.328, q<0.001) and medication (r=-0.298, q<0.001). More medications correlated with decreased HRQOL (VAS, r=-0.298, q<0.001; index r=-0.428, q<0.001). Age correlated to increased medication (r=0.271, q<0.001) and low levels of HRQOL (VAS, r=-0.219, q=0.009; index, r=-0.221, q=0.009).

## Discussion

The main focus of the present study was to explore gut health, nutrient intake and well-being in community-dwelling older adults in order to open up for new therapeutic approaches to promote health and longevity. Our results demonstrate that the prevalence of GI symptoms is as high as 65% in this population. However, our results show a higher prevalence of GI symptoms than previously reported in older populations in other countries (Alameel, Basheikh, & Andrew, 2012; Talley, O’Keefe, Zinsmeister, & Melton, 1992). The difference might be due to variations in the study design and particularly in the methodology chosen for estimating and characterising GI symptoms. It should also be noted that the results are based on self-reported data and rely on the respondents’ honesty, accurate understanding and interpretation of the questions asked. Moreover, it is possible that the present study, focusing on gut health, might have attracted individuals suffering from GI symptoms resulting in a higher prevalence of GI symptoms being reported although any known gastrointestinal disease was exclusion criteria. The majority of the study population was comprised of women (67%), who are known to have more GI symptoms than men (Mapel, Roberts, Overhiser, & Mason, 2013; Peppas, Alexiou, Mourtzoukou, & Falagas, 2008). This could, in addition, skew the results towards a higher prevalence of GI symptoms. Even though we did observe more severe GI symptoms among women we did not detect a higher prevalence of GI symptoms. In the total study population a slightly higher prevalence of dyspepsia and diarrhoea was found compared to previous studies (El-Serag & Talley, 2004; Pilotto et al., 2008). However, the prevalence of constipation, abdominal pain and reflux did correspond to previous findings (Dent, El-Serag, Wallander, & Johansson, 2005; Peppas et al., 2008; Quigley et al., 2006). Hence, indicating that the differences in total gastrointestinal symptoms are most likely due to methodological or cultural dietary variations.

Despite the observation of a generally high level of well-being, our results showed a weak though significant correlation between total GI symptoms and decreased well-being except for depression. For the individual symptom domains, four of the five symptoms depicted by GSRS (diarrhoea, constipation, abdominal pain and dyspepsia) followed the same pattern, while reflux was not found to influence the level of well-being. These results are in accordance with previous reports showing that common GI symptoms and gastrointestinal diseases influence well-being negatively (Barkun & Leontiadis, 2010; El-Serag et al., 2002; Glia & Lindberg, 1997; Kay, Jorgensen, & Schultz-Larsen, 1992; Knowles et al., 2018). The observation that mild GI symptoms correlate to lower levels of well-being among older individuals further emphasise gut health as an important target for improved health.

Dietary intake was self-assessed by a FFQ estimating the intake over the previous year. The FFQ has been validated before (Johansson et al., 2002; Johansson et al., 2010), but only in a Swedish population below the age of 61 years. Hence, for an accurate estimation of dietary intake, a second instrument (i.e. repeated 24 hour dietary recall) could have been used for validation. Using retrospective data collection of dietary intake could lead to recall bias and underreporting (Shim, Oh, & Kim, 2014). The mean self-reported energy intake was found to be low (5.5 MJ/day or 1314 kcal/day) in comparison to the median energy requirement for sedentary individuals (8.5 and 7.1 MJ/day, men and women) according to NNR (*Nordic Nutrition Recommendations 2012* : *integrating nutrition and physical activity*, 2014). The low intake indicates underreporting in the study population, provided body weight balance. Unfortunately we did not have information of body weight or previous change in body weight at the data collection, and therefore we were not able to estimate individual energy requirements. Therefore all macronutrient intakes were reported as E% (e.g. the proportion of total energy intake), to allow for comparisons between participants.

More than half of the study population had an intake of fibre (g/MJ) lower than the NNR, were male participants reported a lower intake than females. A low fibre intake has been associated with constipation as well as inflammatory bowel disease (Ananthakrishnan et al., 2013; Yang, Wang, Zhou, & Xu, 2012). In addition, we recently demonstrated that dietary fibres from yeast have the potential to strengthen the intestinal barrier towards stress-induced intestinal permeability in older individuals suffering from moderate GI symptoms (J. P. Ganda Mall et al., 2018). However, no correlation was found between a low fibre intake and GI symptoms or increased stress-levels, respectively. Our results also demonstrate a high consumption of alcohol as 27% of the study population reported an E% from alcohol that being above recommendations. However, we did not identify any correlation between increased alcohol intake and GI symptoms or well-being.

Our results demonstrate an imbalance in E% fat intake, as energy from saturated fat (E%) exceeded the NNR for as many as 96% of the study population and about half of the population (54%) had and E% from unsaturated fats below the recommendation. An association between saturated fats and constipation in adults (Taba Taba Vakili, Nezami, Shetty, Chetty, & Srinivasan, 2015) have previously been observed, however we did not identify such a relationship in the present study. Nevertheless, our findings suggests that further intervention studies testing a dietary patterns high in protein, fibres and unsaturated fats and less saturated fat might be of benefit for optimising health among older people.

More than half of the study population reported a lower E% from protein than recommended. This could indicate a low protein intake in a majority of the population and particular among men, of whom the majority had a lower E% intake of protein (70%, n=49). This is a substantially higher proportion than reported in a previous study, where only 10% of community-dwelling older people ate less protein than 0.7 g/kg/day, where their study population measured by E% also had higher mean intake than our population (Tieland, Borgonjen-Van den Berg, van Loon, & de Groot, 2012). Interestingly, a low E% energy intake of protein did significantly correlate with symptoms of dyspepsia. In addition, a close but not significant correlation (p=0.055) was observed between intake of estimated added sugar and dyspepsia, though we did see a difference based on group levels where those with GI symptoms had significantly higher intakes of carbohydrates and estimated added sugars than those without GI symptoms. Relationships between dyspepsia and increased total intakes of fat and carbohydrates have previously been observed (Duncanson, Talley, Walker, & Burrows, 2018). Symptoms of dyspepsia did further correlate significantly to decreased well-being. Hence, suggesting that an improvement of protein intake or dyspepsia symptom might increase health (Deutz et al., 2014) and well-being. Moreover, dyspepsia is associated with a higher societal economic burden (Barkun & Leontiadis, 2010) and cost-effective strategies could be useful when the ageing population is increasing. Hence our findings emphasize the importance of an adequate protein intake in older people and calls for targeted intervention studies to thoroughly elucidate the influence of protein intake on dyspepsia or vice versa.

In conclusion, our results demonstrate that the majority of the community-dwelling older adults have a GI symptom, as well as an imbalanced macronutrient dietary intake of saturated fats, fibers and protein, where E% of protein were associated with dyspepsia. Hence, future intervention studies are needed to clarify the causality as well as elucidate specific diets and dietary components as a tool to improve gut health and well-being among community-dwelling older individuals.

## Data Availability

All data is available upon request after publication

